# Detecting SARS-CoV-2 lineages and mutational load in municipal wastewater; a use-case in the metropolitan area of Thessaloniki, Greece

**DOI:** 10.1101/2021.03.17.21252673

**Authors:** Nikolaos Pechlivanis, Maria Tsagiopoulou, Maria Christina Maniou, Anastasis Togkousidis, Evangelia Mouchtaropoulou, Taxiarchis Chassalevris, Serafeim Chaintoutis, Chrysostomos Dovas, Maria Petala, Margaritis Kostoglou, Thodoris Karapantsios, Stamatia Laidou, Elisavet Vlachonikola, Anastasia Chatzidimitriou, Agis Papadopoulos, Nikolaos Papaioannou, Anagnostis Argiriou, Fotis Psomopoulos

## Abstract

The SARS-CoV-2 pandemic represents an unprecedented global crisis necessitating novel approaches for, amongst others, early detection of emerging variants relating to the evolution and spread of the virus. Recently, the detection of SARS-CoV-2 RNA in wastewater has emerged as a useful tool to monitor the prevalence of the virus in the community. Here, we propose a novel methodology, called ***lineagespot***, for the detection of SARS-CoV-2 lineages in wastewater samples using next-generation sequencing. Our proposed method was tested and evaluated using NGS data produced by the sequencing of three wastewater samples from the municipality of Thessaloniki, Greece, covering three distinct time periods. The results showed a clear identification of trends in the presence of SARS-CoV-2 mutations in sewage data, and allowed for a robust inference between the variants evident through our approach and the variants observed in patients from the same area time periods. Lineagespot is an open-source tool, implemented in R, and is freely available on GitHub.

## Introduction

Nearly a year after the first report of SARS-CoV-2 in Wuhan, China, the virus has spread at an unprecedented pace causing a global pandemic. As the main transmission process of the SARS-CoV-2 virus is through droplets and the contact between people, several testing strategies identify whether a person is infected and, in cases of a positive sample, what is the underlying virus variant. However, these methods are not easily scalable, especially in large urban areas. Interestingly, the viral RNA can also be detected in wastewater, with SARS-CoV-2 RNA levels in wastewater correlating with the COVID-19 epidemiology^1–3^. Indeed, in the previous work of Petala et al^3^. normalized viral copy levels in Thessaloniki sewage were in agreement with the epidemiological conditions in the city. Thessaloniki is the second largest city in Greece with around 1 million inhabitants. The city is a chief gateway for entrepreneurs, traders, university students and tourists visiting Greece and, as such, it was the place where the Greek patient “zero” appeared in March 2020. Thessaloniki is also the place where the so-called South African mutation appeared in Greece for the first time. In other words, identifying new variants in the city sewage is critical to understand further scattering to the rest of the country.

The presence of SARS CoV-2 RNA in sewage provides us with a unique opportunity, i.e. to identify the most prevalent virus lineages through the analysis of the traces evident in wastewater samples. So far, although there are few studies exploring the SARS-CoV-2 diversity in sewage, it still remains an open issue as there are no widely accepted methods that can sufficient address this. The most common used approaches involve the sequencing the wastewater samples, and the consequent application of low frequency variant analysis methods^4^ or metagenomic approaches^5,6^. In either case, the interpretation of the results focuses on the detection of specific variants^4^ or lineages^2^ such as B.1.1.7 and 501.V2, prevalent clades^6^ (19A, 20A and 20B) or new uncharacterized mutations^6^.

In this work we propose a novel methodology called ***lineagespot***, implemented as a software tool, that can facilitate the detection of SARS-CoV-2 lineages in wastewater samples using next generation sequencing. The method is tested and validated across three municipal wastewater samples retrieved in Thessaloniki, Greece in three different time periods, and correlated with the lineages observed in patients from the same area time points. Based on a variation of the Illumina Arctic pipeline for the identification of mutations at low frequencies (< 0.01), and the lineage assignments defined by Pangolin, this method identifies all SARS-CoV-2 mutations present in the wastewater of Thessaloniki, and attempts to infer the potential distribution of the SARS-CoV-2 lineages. The methodology is proven to be effective in detecting the mutational load in the wastewater, with the inferred lineages being roughly aligned to the predominant lineages identified through targeted (and therefore biased) patient-derived genotypes.

## Results

### Comparison of variant calling methods

The proposed methodology was applied on a selected sewage sample (corresponding to the 05-11 February 2021 time period), and for which three different variant callers were assessed: 1) *freebayes, 2) mpileup* and 3) *GATK Mutect2* (cancer only mode). In terms of parameters, *freebayes* was used with a low variant frequency parameter of 0.01, *mpileup* reported every position (either reference, or variant), and *GATK Mutect2* was used with the default parameters.

An example of the output produced by the methodology, regardless of the variant calling method, is shown in **Table 1**. In this table the overlap between the pangolin’s rules and the rules generated by the tool for the input dataset is captured for each lineage. In order to quantify the overlap, three basic metrics are produced; the overlap by considering pangolin’s rules as a decision tree (*Tree Overlap*), the total overlap regardless of the rules order (*Total Overlap*), and the overlap for the rules that are satisfied only by the identified mutations (i.e. explicitly listed in the variants’ file), and therefore excluding all rules based on the unmutated reference (*Total Overlap Var*). In addition to the previous metrics, the related ratio values are also calculated (*Tree ratio, Total Ratio, Total Ratio Var*). Finally, information regarding the read depth for each position (reference and variant) is also provided in the output file.

**Table 1:**
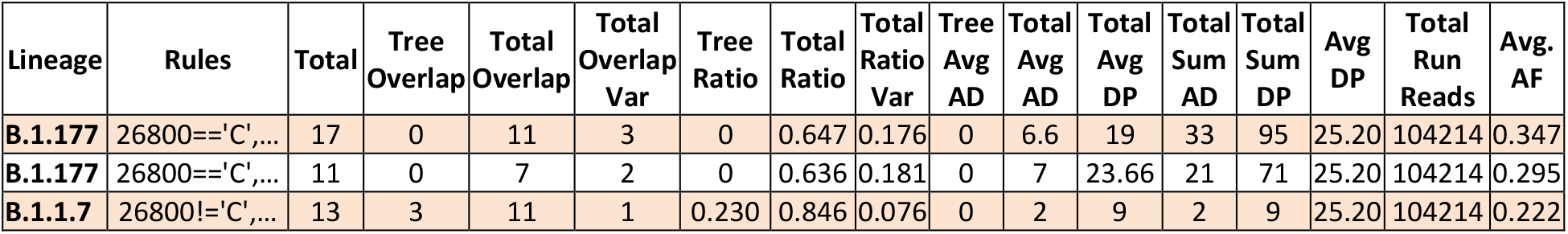
Each row in the table corresponds to a single lineage rule defined by pangolin. The columns correspond to the different metrics captured, in order to perform the systematic evaluation.

Based on this detailed table, a second output is generated as a simplified summary. In this case, all rows for which the *Total Overlap Var* column was equal to 0 were removed, and therefore potential lineages that would be assigned based only on the unmutated reference (i.e. no actual mutations detected) are excluded from the analysis. The remaining rows were collapsed (**Table 2**) through a process in which the average values of the basic metrics were calculated for each lineage; i.e. the mean of the *Tree Ratio*, the *Total Ratio*, the *Total Ratio Var*, the *Tree Av AD*, the *Total Av AD*, and the *Avg AF* columns.

**Table 2:** Each row in the table corresponds to a unique lineage, after the merging process. The remaining metrics can be consequently used to assess the presence of the particular variant in the dataset.

| Lineage | Mean Tree Ratio | Mean Total Ratio | Mean Total Ratio Var | Mean Total Av AD | Mean AF |
| --- | --- | --- | --- | --- | --- |
| B.1.1.7 | 0.230769231 | 0.846153846 | 0.07692307 | 2 | 0.222222222 |
| B.1.177 | 0.002222635 | 0.610840685 | 0.05660585 | 9.76863806 | 0.488594472 |

Depending on the variant caller tool used (*freebayes, mpileup* and *GATK Mutect2)*, ***lineagespot*** generates a unique output. All outputs are compared pairwise, based on the decision tree rules (*n*_*d*_), and the total number of rules satisfied (*n*_*t*_). For each lineage, the absolute values of the differences between the two metrics (*n*_*d*_, *n*_*t*_) of the files are calculated. As an example, for lineage *A*.*1*, the output produced by using the *freebayes* variant caller tool in the first step of the methodology, returns *n*_*d*_ = 2 and *n*_*t*_ = 10 rules satisfied, while the output of the *GATK Mutect2* tool returns *n*_*d*_ = 1 and *n*_*t*_ = 4 rules satisfied. As a result, the two outputs exhibit a difference of *n*_*d*_ = 1 and *n*_*t*_ = 6 rules (**Table 3**).

**Table 3:** Snapshot of the difference between the two metrics (*n*_*d*_, *n*_*t*_) between the output files coming from *freebayes* variant caller and *mpileup*.

| Lineage | Rules | $N_t$ | $n_d^{\text{freebayes}}$ | $n_t^{\text{freebayes}}$ | $n_d^{\text{mpileup}}$ | $n_t^{\text{mpileup}}$ | $\vert n_d^{\text{freebayes}} - n_d^{\text{mpileup}}\vert $ | $\vert n_t^{\text{freebayes}} - n_t^{\text{mpileup}}\vert $ |
| --- | --- | --- | --- | --- | --- | --- | --- | --- |
| B.1.1.7 | 26800!= 'C', ... | 13 | 3 | 72 | 3 | 71 | 0 | 1 |
| B.1.177 | 26800== 'C', ... | 66 | 3 | 72 | 3 | 69 | 0 | 3 |
| B.1.177 | 26800== 'C', ... | 6 | 3 | 72 | 3 | 69 | 0 | 3 |
| B.1.351 | 26800!= 'C', ... | 38 | 3 | 162 | 3 | 160 | 0 | 2 |

In addition, the total number of lineages are shown in **Table 4**. In the same table, the maximum absolute *n*_*d*_ difference and the maximum *n*_*t*_ difference for each pair of files are calculated. The latter is used for an overall comparison of the output files.

**Table 4:** Summary table of the three output files produced by *freebayes, mpileup* and *gatk mutect2* variant caller. The three files are compared in pairs.

| Files | Number of differences | max absolute $n_d$ difference | max absolute $n_t$ difference |
| --- | --- | --- | --- |
| freebayes - gatk | 3791 | 4 | 31 |
| freebayes - mpileup | 3140 | 4 | 33 |
| gatk - mpileup | 1571 | 1 | 31 |

Based on the above comparison, we consider that the most productive and informative approach is to utilize *freebayes* as the variant calling tool. The rest of the results shown below, are based only on the *freebayes* tool output.

### Evaluating lineage-specific mutations across time periods

The proposed methodology was applied on three sewage samples, across three time periods: 02-14/12/2020 (*SampleA*), 05-11/02/2021 (*SampleB*) and 12-18/02/2021 (*SampleC*).

**Table 5:** The number of reads of each sample across the different stages of the applied bioinformatics analysis.

|  | SampleA | SampleB | SampleC |
| --- | --- | --- | --- |
| Total number of reads in raw fastq files | 176380 | 466734 | 1567696 |
| Total number of reads after adaptor trimming | 171312 | 453278 | 1565826 |
| Total number of mapped reads | 77444 | 133073 | 836105 |
| Total number of sorted mapped reads | 77105 | 131744 | 834085 |
| Total number of input reads to ivar | 75396 | 128013 | 811508 |
| Total number of ivar shortened reads | 4013 | 15853 | 37245 |
| Reads outside primer region | 1194 | 1937 | 12178 |
| Total number of reads after primer trimming | 71898 | 113954 | 784662 |
| Sorted reads after primer trimming | 69706 | 104214 | 769744 |
| Mapped sorted reads after primer trimming (duplicates removal) | 69706 | 104214 | 768820 |

**Figure 1:**
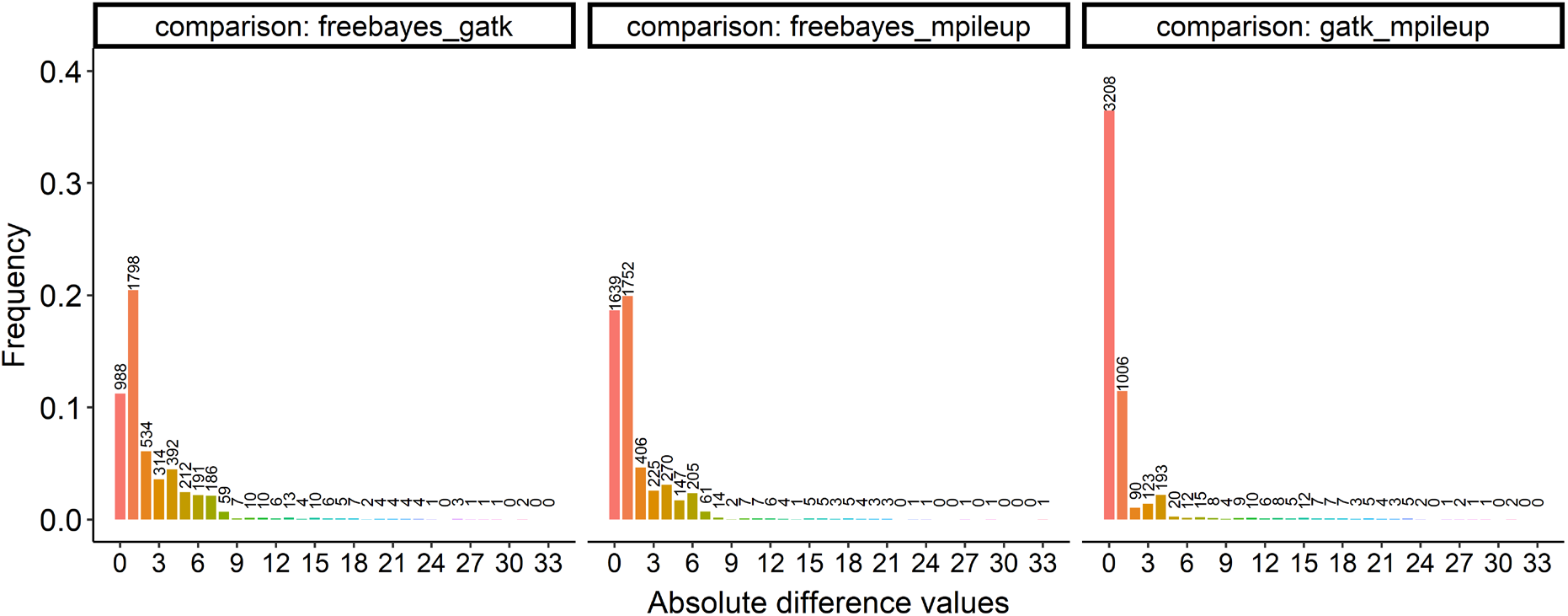
Distribution of the absolute *n*_*t*_ difference values between the output of the three variant calling tools use (pairwise comparisons).

**Figure 2:**
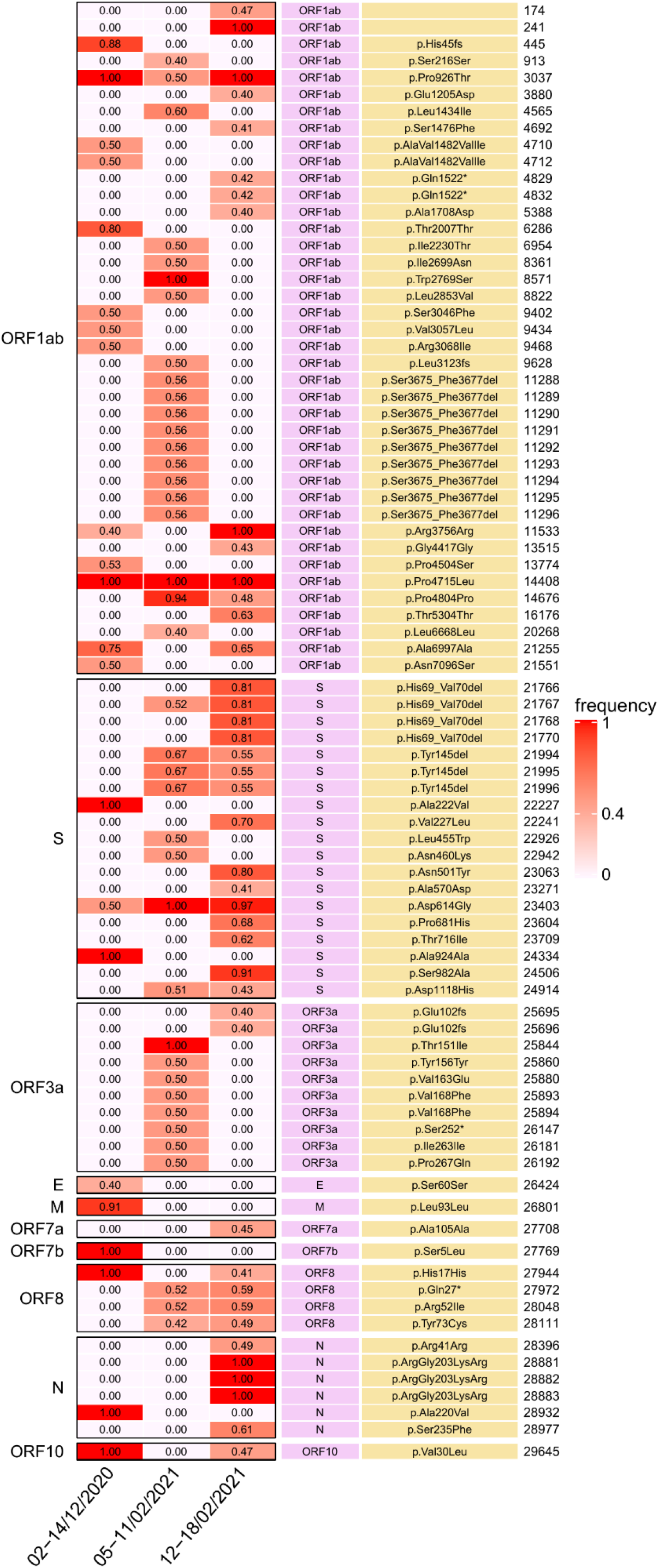
Mutation positions with allele frequency (AF) > 0.4.

**Figure 3:**
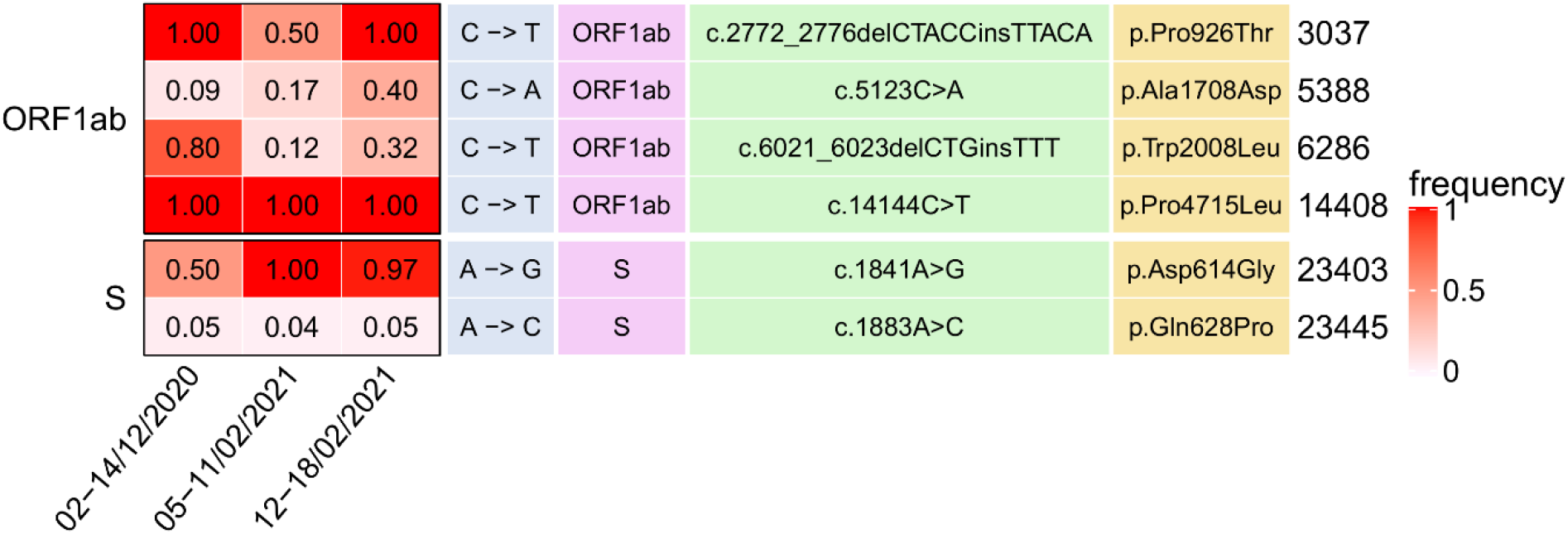
Common mutation positions across all time points.

**Figure 4:**
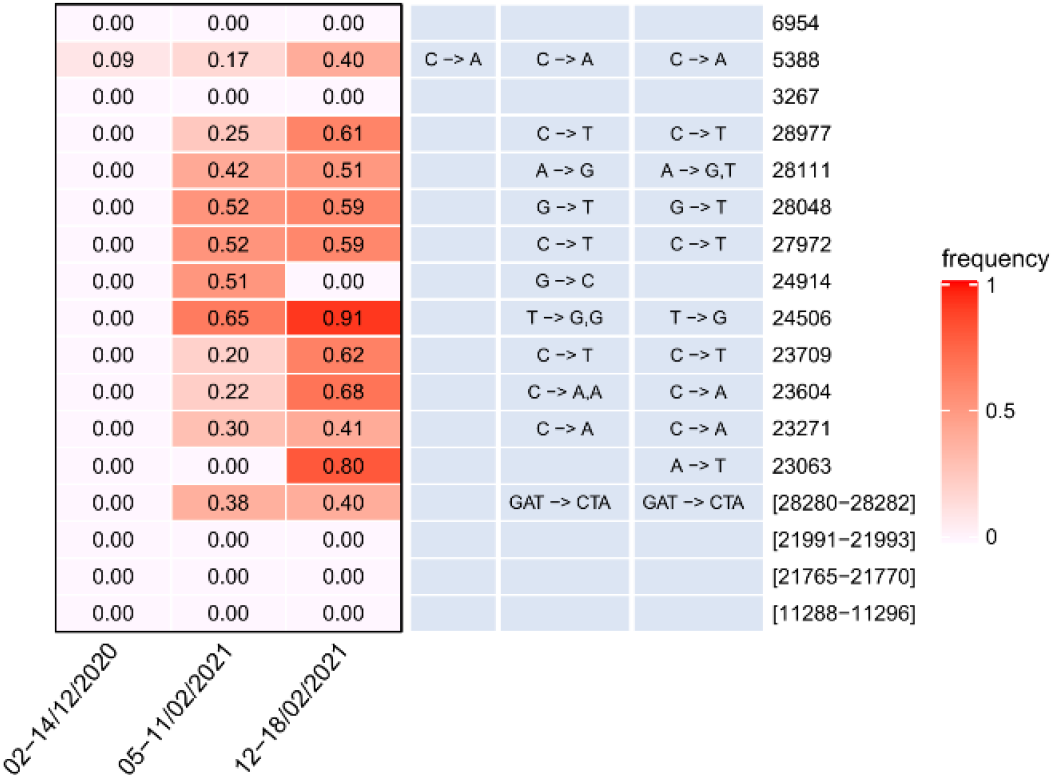
Detected B.1.1.7-detected mutations (UK Lineage)

**Figure 5:**
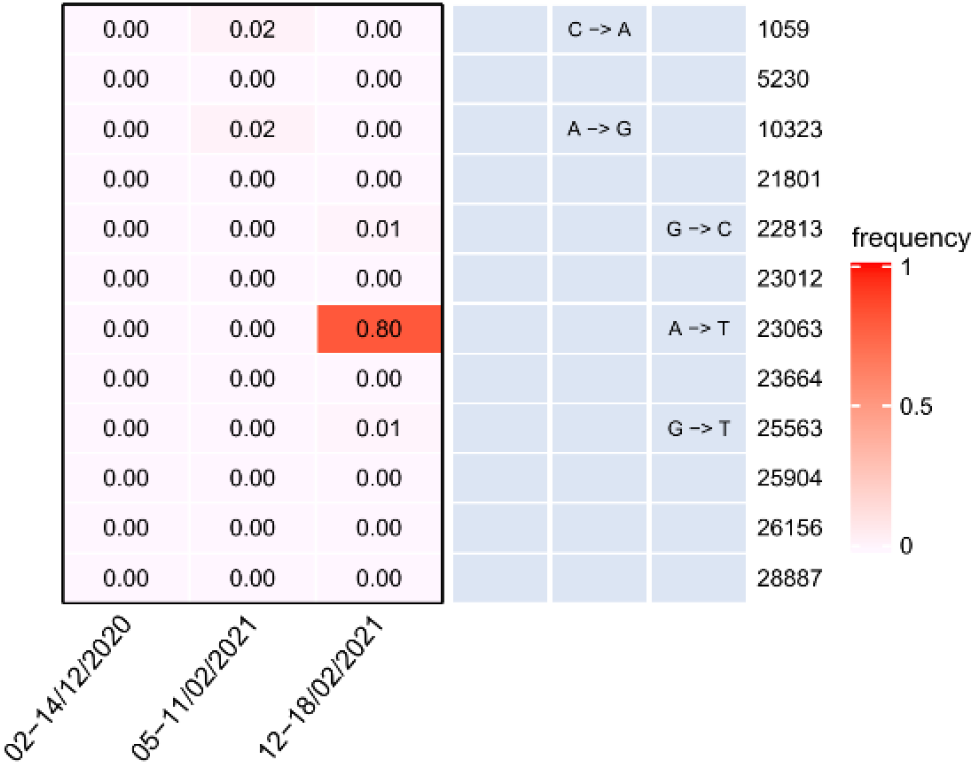
Detected B.1.351-detected mutations (South Africa lineage)

### Assessing lineage assignment

Qualitative assessment between the major lineages found in the biased sampling and the three time periods.

**Period 1**: 02-14/12/2020

In this time period, the most frequent lineage detected in targeted sampling was B.1.177 (on average, 80% of the samples captured). The application of our method to the sewage sample of the same period, detected the same lineage (B.1.177) with the following characteristics:

| Lineage | Mean Tree Ratio | Mean Total Ratio | Mean Total Ratio Var | Mean Total Av AD | Mean AF |
| --- | --- | --- | --- | --- | --- |
| B.1.177 | 0.002246535 | 0.605276332 | 0.042082807 | 5.626164875 | 0.513229318 |

**Period 2**: 05-11/02/2021

In this time period, the two most frequent lineages detected in targeted sampling were B.1.177 (~56% of the samples captured) and B.1.1.7 (~35% of the samples captured). The application of our method to the sewage sample of the same period, detected the same lineages (B.1.177 and B.1.1.7) with the following characteristics:

| Lineage | Mean Tree Ratio | Mean Total Ratio | Mean Total Ratio Var | Mean Total Av AD | Mean AF |
| --- | --- | --- | --- | --- | --- |
| B.1.1.7 | 0.230769231 | 0.846153846 | 0.076923077 | 2 | 0.222222222 |
| B.1.177 | 0.002222635 | 0.610840685 | 0.05660585 | 9.76863806 | 0.488594472 |

**Period 3**: 12-18/02/2021

In this time period, the two most frequent lineages detected in targeted sampling were B.1.177 (~54% of the samples captured) and B.1.1.7 (~38% of the samples captured). The application of our method to the sewage sample of the same period, detected the same lineages (B.1.177 and B.1.1.7) with the following characteristics:

| Lineage | Mean Tree Ratio | Mean Total Ratio | Mean Total Ratio Var | Mean Total Av AD | Mean AF |
| --- | --- | --- | --- | --- | --- |
| B.1.1.7 | 0.230769231 | 0.846153846 | 0.153846154 | 31.33333333 | 0.5875 |
| B.1.177 | 0.006394285 | 0.627302601 | 0.029552992 | 23.74139477 | 0.500008793 |

## Discussion

Analyzing wastewater — used water that goes through the drainage system to a treatment facility — is a way that researchers and surveillance systems can track pathogens that are excreted in urine or feces, such as SARS-CoV-2. Monitoring effluents could provide better estimates on coronavirus spread than sampling and testing the population, because wastewater surveillance can account for those who have not been tested and have only mild or no symptoms. Moreover, an effective and reliable methodology able to detect virus load and SARS-CoV2 variants from municipal wastewater samples could drastically, or at least help, decrease the cost of virus variant detection in the general population based on whole genome sequencing.

In this manuscript we present and validate a methodology named ***lineagespot***, making use of Next Generation Sequencing data, able to detect lineages and mutational load of SARS-CoV2. The methodology aims to aid the epidemiological system for the monitoring of COVID 19 pandemic in urban areas.

The method has been tested in different time point samples taken from the main Municipal Wastewater Treatment Plant of Thessaloniki - Greece, where effluents from approx. 750.000 inhabitants are collected. The ***lineagespot*** method demonstrated to be sensitive enough to identify and quantify differences in the mutational load, across various time points. Moreover, the quantitative data obtained using ***lineagespot*** are in accordance with the trends of well-known mutations (such as Asp614Gly) in the same period with the overall epidemiological status of the municipal area. The application of ***lineagespot*** in such complex samples, like those from Wastewater Treatment Plants, was able to assign lineages and in agreement with the trend of the major lineages detected in the area of Thessaloniki, in the three time points by whole genome sequencing of samples from the general population.

Overall, the method compared to other ones (Sanger sequencing)^4,7^ resulted more informative, sensitive enough to detect mutations with low frequency and able to assign with good approximation the correct lineage present in the municipality.

## Methods

### Sampling and isolation

Wastewater samples were collected from the entrance of the main Municipal Wastewater Treatment Plant of the city which accommodates sewerage of about 750.000 inhabitants. Wastewater entering this Plant refers exclusively to citizens from urban districts of the city. Typical values of certain physicochemical properties of wastewater samples tested in this study are displayed in **Table 1**. These properties demonstrate, among others, the existence of suspended solids, dissolved organic matter, dissolved oxygen and salinity that may have strong impact on viral adsorption and decay because of oxidation and increased metabolic activity of microorganisms in sewage. The residence time of sewage until the entrance of the Plant is between 2 and 7 hours (depending on the area), which is more than enough to allow viral adsorption and decay. Identification of mutations may be hindered by viral adsorption and decay and for this reason the present effort is particularly significant.

**Table 1.** Main quality characteristics of wastewater samples

| Parameter/ Sample period | 02-14/12/2020 | 05-11/02/2021 | 12-18/02/2021 |
| --- | --- | --- | --- |
| pH | 7.5 ± 0.15 | 7.8 ± 0.10 | 7.8 ± 0.15 |
| Electrical Conductivity (S/cm) | 8.5 ± 1.5 | 9.6 ± 2.7 | 4.6 ± 1.0 |
| Total Suspended Solids (mg/L) | 620 ± 80 | 930 ± 115 | 1200 ± 125 |
| BOD<sub>5</sub> (mg/L) | 385 ± 75 | 525 ± 40 | 650 ± 65 |
| COD (mg/L) | 960 ± 120 | 1250 ± 150 | 1570 ± 170 |
| Dissolved Organic Carbon (mg/L) | 35.0 ± 6.5 | 49.0 ± 10.5 | 44.0 ± 5.0 |
| UV absorption at 254 nm (1/cm) | 0.35 ± 0.05 | 0.40 ± 0.06 | 0.45 ± 0.05 |
| Total Nitrogen (mg/L) | 62.0 ± 15.0 | 76.0 ± 11.5 | 95.0 ± 12.0 |
| Ammonium Nitrogen (mg/L) | 28.5 ± 8.0 | 33.0 ± 3.0 | 38.0 ± 4.0 |
| Total Phosphorus (mg/L) | 11.5 ± 4.5 | 11.5 ± 1.5 | 12.0 ± 2.0 |

Sampling and handling of the sewage samples were performed according to Petala et al^3^. Briefly, samples were obtained using a refrigerated autosampler (6712 Teledyne ISCO) programmed to deliver a 24-hours composite sample by mixing consecutive half-hour samples. Samples were transported to the lab on ice and were processed immediately. Three 50-mL aliquots of each untreated wastewater sample were subjected to centrifugation at 4000xg for 30 min. Afterwards, a composite sample was obtained from supernatants and pH was adjusted to 4 using 2 M HCl solution. Then, three aliquots of 40 mL each, were filtered through 0.45-μm pore-size, 47-mm diameter electronegative membranes (HAWP04700; Merck Millipore, Ireland) followed by RNA extraction as described in Ahmed et al^8^.

### RNA extraction and SARS-CoV2 detection

Extracted RNAs originating from 12 processed electronegative membranes and spanning different days of sampling were filtered through OneStep PCR inhibitor removal kit (Zymoresearch) according the manufacturer’ s instructions, pooled and concentrated using the NucleoSpin® RNA XS, Micro kit (MACHEREY-NAGEL GmbH & Co. KG, Düren, Germany). Concentrated RNAs were subjected to real-time RT-PCR testing for SARS-CoV-2 quantification, utilizing the N2 protocol proposed by the Centers for Disease Control and Prevention (CDC) for the diagnosis of COVID-19 in humans (CDC, 2020). The assay was performed on a CFX96 Touch™ Real-Time PCR Detection System (Bio-Rad Laboratories, Hercules, CA, USA). A calibration curve was generated and SARS-CoV-2 viral loads were expressed as genome copies per μl of RNA extract. In total, three concentrated RNA pools from respective time periods were quantified; 02-14/12/2020 (36 copies/μl), 05-11/02/2021 (68 copies/μl), and 12-18/02/2021 (53 copies/μl).

### Library preparation and sequencing

The targeted sequencing method was applied by preparing 400nt amplicons using the ARTIC v3 protocol developed by Wellcome Sanger Institute^9^, with some modifications. First, cDNA synthesis was prepared from 10 ul of RNA using SuperScript II reverse transcriptase (Invitrogen - Thermo Fisher Scientific, USA) and 50 ng/ul of random primers according to the protocol guidelines. For subsequent cDNA amplification, 2.5 ul of the generated cDNA was used instead of 6 ul, using ARTIC PCR primer pools (v3). Finally, the NEBNext adaptor (New England Biolabs, US, #7335) was used in the ligation reaction, diluted with adaptor dilution buffer at 10μM final concentration. All purification steps were performed according to the ARTIC protocol. The samples were paired-end sequenced on a MiSeq platform (Illumina, USA) with a read length of 2 × 300 bp.

### Raw data analysis

The initial phase of the bioinformatics analysis is to produce an alignment of the sequencing reads, while maintaining extremely strict criteria, in order to remove any potential contaminants and/or sequencing errors. The first step is the adaptor removal process, where any adaptors are removed from the raw *fastq* sequences, with the cleaned reads mapped to the SARS-CoV-2 reference genome (Wuhan variant, NC_045512), using minimap2 tool^10^ with a minimal chaining score (matching bases minus log gap penalty) equal to 40. From this process, only the paired-end sequences are retained, while any other (unmatched, multiple mappings, etc.) are removed. In the next step, the primer sequences are excluded using the *ivar* tool^11^, setting a minimum of 200 length in nucleotides for a read to be retained after trimming, and a minimum threshold for sliding window of 15 quality to pass (width of sliding window equal to 4). The final sequences are then remapped to the same reference genome (minimal chaining score equal to 40). Finally, and in order to be able to detect low frequency variants, the *freebayes* variant caller was used with a low variant frequency parameter of 0.01. Ultimately, all identified mutations were annotated using the *SnpEff* tool^12^ and the NC_045512.2 (version 5.0) database.

### Downstream analysis of lineages detection

In order to identify and assign different SARS-CoV-2 lineages based on the mutations detected from a single sewage sample, we implemented the proposed methodology in a tool named ***lineagespot***. The tool accepts as input a VCF file, which contains all mutations identified in the sample, along with the reference SARS-CoV-2 genome file, and a file containing all lineage-assignment rules, as retrieved from the [pangolin](https://github.com/cov-lineages/pangolin) tool^13^ repository. After analyzing all inputs, a tab-delimited file (TSV file) is produced containing the most probable lineages that have been found. Figure 1 shows an overview of the tool’s functionalities, which can be described in 2 phases:

#### i) Creating rules from variants

In this phase all rules that can be derived from the VCF file are constructed. Initially, a vector of all genome positions is created, for which each position is set to be equal to the reference genome nucleotides. Then, the VCF file is read and a set of new rules is formed by setting each position of the file with the reported variant or multiple variants (in case there is more than one reported variant at the same position). It should be noted that positions that have been detected with more than one variant, should include all of them at the VCF’s ALT column in a comma-delimited format. Most of the variant caller tools (*freebayes, GATK*, etc.) are doing this by default. Finally, positions with reference read depth equal to zero are removed from the first vector. The remaining two vectors are merged into one.

In addition to finding all positions that need to be set equal to the base that has been allocated, four more rules are added for each genome position. These rules contain all bases *not* equal to the nucleotide of the original rule. For example, if position 5388 is equal to base ‘A’ (representing rule *5388==‘A’*), then four new rules are added containing all bases not equal to ‘A’, e.g., *5888!=‘T’, 5388!=‘G’, 5388!=‘C’, 5388!=‘-’* (where the *‘-’* symbol stands for a gap in the referred sequence). Finally, all rules are merged into a single vector representing this particular lineage.

**Figure 6:**
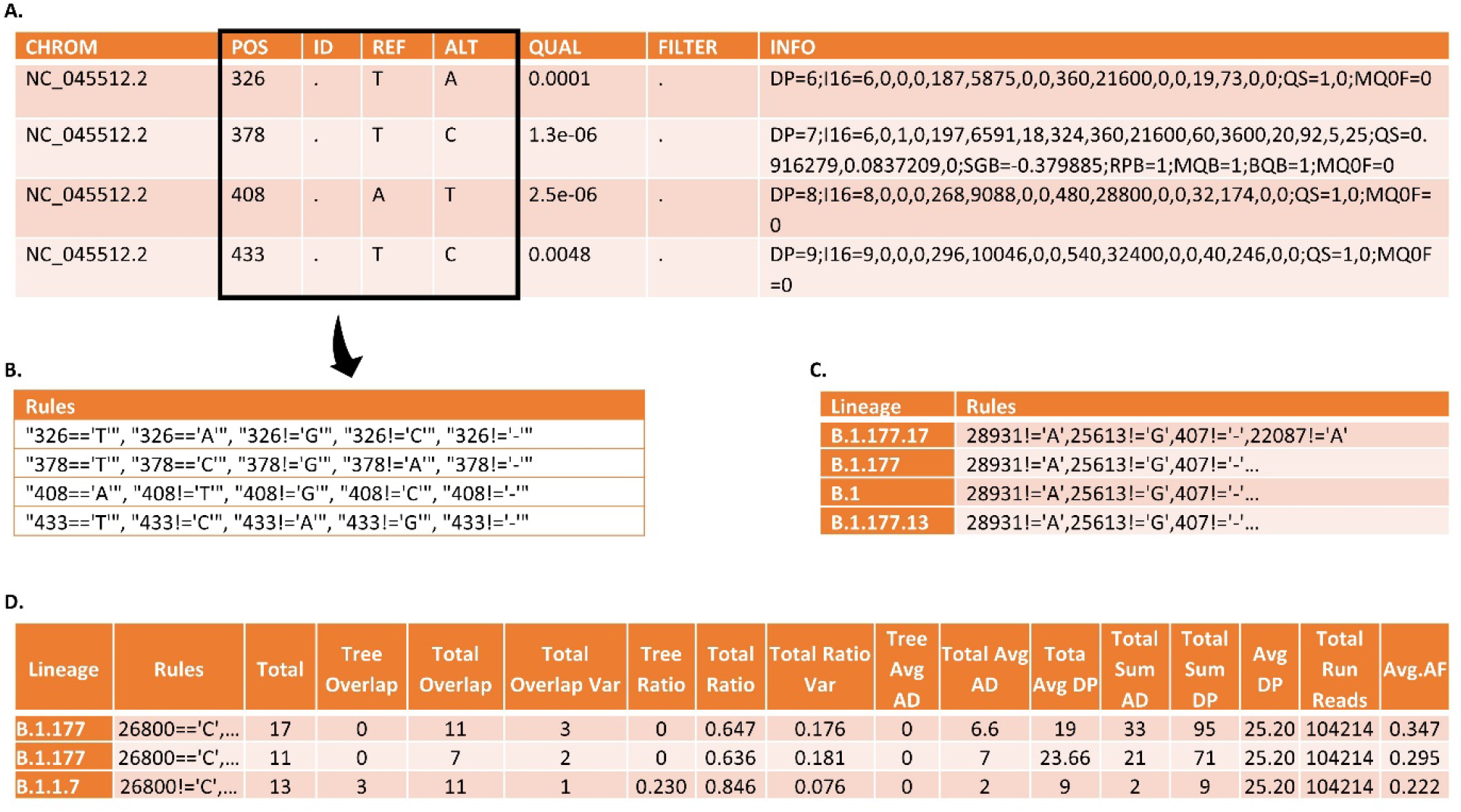
Snapshots of the intermediate steps. **A**. A VCF file produced by the chosen variant caller. **B**. The rules as they are generated by the VCF file. **C**. Pangolin’s decision rules. **D**. A tab-delimited file as produced by ***lineagespot***.

#### ii) Comparing with pangolin rules

The second phase aims to compare the rules derived from the VCF file with the assignment rules provided by the pangolin tool. Specifically, all decision rules are read from the input pangolin file, and for each lineage, the related rules are compared with the final rule vector.

Three metrics are computed and stored in the output file; the total number of rules leading to the related lineage (*N*_*r*_), the number of rules satisfied by the created rule vector, considering pangolin’s rules as a decision tree (*n*_*d*_), and the total number of rules satisfied (*n*_*t*_). Also, the related ratio values are being computed, giving a satisfaction percentage of each lineage:

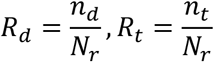

### Underlying assumptions of the method

It should be noted that the methodology relies on the following assumption. Given a group of reads that satisfy a rule A of lineage L, and another group of reads that satisfy rule B from the same lineage L, then the lineage L is incorrectly assigned. As an example, suppose that a group of reads satisfy only the first two rules from lineage *B*.*1*.*177*.*17* (*28931!=‘A’, 25613!=‘G’*), and another group of reads satisfy the next two rules from the same lineage (*407!=‘-’, 22087!=‘A’*). Based on the method description above, lineage B.1.177.17 will be marked as an identified lineage, even though none of the reads satisfy all the rules of the lineage.

In order to mitigate this risk, we are taking into consideration a number of different indicators, that reflect the number of total rules satisfied, the number of rules that are satisfied based on the detected mutations, and the overall number of reads that support both reference and allele for each of the rules.

## Data Availability

The implemented code that produces the results of this paper, starting from the VCF files, is available on the GitHub repository: https://github.com/BiodataAnalysisGroup/lineagespot.

The raw FASTQ files are available through Zenodo: https://zenodo.org/record/4564182

https://doi.org/10.5281/zenodo.4564182

## Acknowledgements / CRediT author statement

This work was supported by the “*Greece vs Corona: Flagship Action to address the SARS-CoV-2 crisis. Epidemiological study in Greece through extensive testing for virus and antibody detection, viral genome sequencing and genetic analysis of patients*” project, which is funded by the General Secretariat for Research and Innovation, under the Public Investments Program (PIP). Additionally, this work was supported by *ELIXIR*, the research infrastructure for life-science data.

NP, MT, MCM and AT developed the ***lineagespot*** code and performed the downstream analysis. NP and MT performed the analysis of the raw NGS data. EM, SL and EV did the library preparation. CD and AC provided the data and reviewed the submitted version. MP and MK participated in experimental investigation. AP, NP, and TK participated in project conceptualization, management and funding. FP and AA designed, supervised the study, and reviewed the manuscript. All authors contributed to the article and approved the submitted version.

## Supplementary Material

### S1: Code and data

The raw FASTQ files are available through Zenodo: https://zenodo.org/record/4564182

